# Assisted Reproductive Technology Dataset of Embryo Time-lapse Images and Clinical Data

**DOI:** 10.1101/2024.11.01.24316563

**Authors:** Dmytro Zhylko, Raquel Del Gallego, Sarah Pardo, Rameen Mahmood, Ya Tung Hsieh, Salma Selim, Daniela Nogueira, Ibrahim El-Khatib, Barbara Lawrenz, Human M. Fatemi, Farah E. Shamout

## Abstract

In this report, we present Version 1.0 of the Assisted Reproductive Technology (ART) Dataset, a multi-modal fertility dataset from treatments performed at the ART Fertility Clinic in Abu Dhabi, United Arab Emirates, between 2015 and 2022. The data consists of Electronic Health Records (EHR) and embryo development image sequences captured with the Vitrolife EmbryoScope time-lapse system, providing detailed treatment, morphology, and pregnancy outcome information. The final processed dataset consists of a total of 14,776 embryos from 1,810 patients across 2,500 treatments. This dataset will be used in the development of machine learning models for automated analysis of embryo development and viability, to assist clinical decision-making. This report provides a summary of the statistics of the dataset, as well as the extraction and pre-processing pipelines of the time-lapse images and EHR data. The dataset is private, so we publish this report for transparency on data pre-processing pipelines to share the methodology with similar studies that may arise.

## 1. Introduction

Assisted Reproductive Technology (ART) refers to medical treatments and procedures designed to assist individuals experiencing difficulties in achieving spontaneous pregnancy conception. Infertility is estimated to affect 15% of the global population, over 48 million couples worldwide (1, 2). However, each treatment cycle has a high financial, physical, and emotional cost. This motivates ongoing research to achieve higher success rates by optimizing treatment protocols. Embryo selection is a critical step in ART cycles, and early, accurate assessment of embryo viability can dramatically impact treatment outcomes. Historically, embryo evaluation has been performed through microscopic assessment at specific time points, disturbing the embryo’s controlled incubation environment. Thanks to the introduction of time-lapse imaging systems, designed with built-in cameras, embryologists have been able to monitor development with unaltered conditions. Despite the controversy surrounding whether time-lapse technology has a real impact on clinical outcomes (3), the high-resolution data produced is key for use in developing computational learning models.

In this report, we present the Assisted Reproductive Technology (ART) Dataset, which contains data from 14,776 embryos cultured at the ART Fertility Clinic in Abu Dhabi, United Arab Emirates, between 2015 and 2022. Each embryo has a time-lapse image sequence captured during its development in the Vitrolife EmbryoScope time-lapse system^∗^, combined with Electronic Health Records (EHR) of information about the patient and treatment protocol. In Section 2, we present a summary of the contents of the final processed dataset and provide the statistics of its features and labels. In Section 3, we describe the pre-processing pipelines used to extract and clean the raw images and records sourced from the clinic.

## 2. Overview

### 2.A. Overall statistics

The final dataset contains 14,776 multimodal samples of combined embryo EHR and time-lapse image sequences, collected from 1,810 patients across 2,500 treatment cycles. A treatment cycle refers to a single round of oocyte pick-up and artificial fertilization, as a consequence of the administration of a supplementary hormonal treatment to stimulate ovarian oocyte growth. Patient age ranges from 19 to 53, with an average age of 34.7. The average number of treatment cycles per patient is 1.3 (min. 1, max. 17), and the average number of embryos per treatment cycle is 5.9 (min. 1, max. 12). 13,816 embryos were fertilized from fresh oocytes, and 960 were from frozen oocytes. 13,388 were fertilized using Intra-Cytoplasmic Sperm Injection (ICSI), 91 were fertilized using next-day ICSI, while 1,297 were fertilized using traditional in-vitro fertilization (IVF).
Since next-day ICSI is not a standard practice in every clinic, we introduce it briefly. In next-day ICSI, initially immature oocytes develop to mature oocytes after one day in culture, and then they are fertilized with the ICSI technique. We further analyze cohort characteristics and provide statistics and descriptions in Table 8. We derive several datasets from the initial extraction, following the tasks defined in Section 2.B, and provide sub-dataset sizes in units of single embryo in Tables 3, 2, 5, 6.

### 2.B. Datasets and labels

In our final labeled datasets, multiple labels are associated with each embryo, allowing for a variety of learning tasks. Clinic protocols determine the type of labels and properties which are available in a given dataset. The main outcomes are described in Table 1. We present the statistics and descriptions of the primary label sets of the ART Embryo Dataset: sequence-level blastocyst development labels (Section 2.B.1), image-level development stage labels (Section 2.B.2), sequence-level genetic labels based on biopsy results (Section 2.B.3), and sequence-level pregnancy outcomes (Section 2.B.4).

For the development of deep neural networks, we group the embryo sequences based on the patient identifier, and assign each patient randomly to either the training (60%), validation (20%), or test (20%) set, ensuring that all embryos from a unique patient are present in only one split. We then filter each data split for each task to ensure no overlap across training and test sets across all tasks. This limits amount of patient specific learned features (like patient embedding) we can use, since test time patients are not present in training set, but prevents any potential data leak and tests model robustness to potential distribution shifts.

#### 2.B.1. Sequence-level blastocyst formation

Blastocyst development is subdivided into several stages: start of blastulation (tSB), blastocyst formation (tB), expanded blastocyst (tEB), and hatched blastocyst (tHB). Fig. 2 shows examples of each step in blastocyst development. We considered the presence of any of these labels as a positive blastocyst formation. We assign a positive label to all sequences which have at least one blastocyst label assigned to any image, in case no such label is present, or labels are missing entirely, we cross reference image annotations with EHR data, that contains less accurate indication, like day blastocyst stage was reached. If any of the aformentioned conditions are meet we assign positive label to this sequence, and negative otherwise. While in some cases an embryo may develop to blastocyst but degenerate before it can be preserved or transferred, we consider these to be positive labels for the blastocyst prediction task, and we trim the sequence up to the positive blastocyst stage or day before degeneration, depending on which one is present. The distributions of positive and negative samples in the training, validation and test sets are provided in Table 2. Dataset contains 7,043,561 images, across 14,776 embryo sequences, collected from 1,810 different patients, and is the biggest dataset we extracted.

#### 2.B.2. Image-level development stage

The EmbryoScope system enables embryologists to label images in a sequence with the observed stage of development, with a total of 18 possible stages. The definition of each development stage is provided in Table 3. Our annotations contain cell division stage annotations up to 9 cells, followed by the compaction and blastulation stages. The final dataset contains 6,677,028 labeled images from 1,798 embryo sequences, collected from 1,798 patients. We filter sequences where we suspect incomplete annotation might have occurred, as a result this dataset contains only 17 stages, as we identified sequences with tDead stage as sequences with risk of incomplete annotation. For the full explanation of filtering rules refer to Section 4.A. Table 3 shows the label distributions for training, validation, and test sets. No monotonicity in distributions indicate, that not all of the stage annotations are present in sequence, even if sequence progressed past certain stage. This is the result of interval-based image acquisition, that results in certain stages happening in between captures, especially ones that happen fast, e.g. odd cell count stages.

Additionally, labels might have been missed, due to later well insertion during IVF or next day ICSI protocols, resulting in missed first stages. Considering practical aspect of this dataset, we in Table 4 also report distributions in the sub-sets after label filling, realized as forward filling of the annotations, by assigning previous label to current frame, unless label was already assigned in annotation. We can observe that some stages last significantly shorter then others, which contributes to unbalanced training problem. Fig. 1 shows an example of several embryo images with corresponding label.

#### 2.B.3. Sequence-level ploidy outcomes

Our dataset includes results from PGT for aneuploidy (PGT-A), which detects the number of chromosomes in the embryo cell nuclei (ploidy) to screen for chromosome abnormalities. The ART Dataset uses positive (euploid) and negative (aneuploid) PGT-A results to assign genetic outcome labels to each embryo sequence. In the final dataset, of the 7,872 embryos present in subset from Section 2.B.1, which additionally underwent conclusive genetic testing, 3,653 embryos were identified as euploid, where cells in the biopsy sample contain 23 chromosome pairs, and 4,219 embryos were identified as aneuploid, where the cells contain an abnormal number of chromosomes. Embryos with inconclusive test result were removed from dataset (not included in above numbers). The aneuploid embryos are further categorized by aneuploidy type: complex 1,824 (effects more then one chromosome), segmental 790 (gain/loss of chromosome segment), mosaic 336 (MVA syndrome), whole chromosome 3,495 (gain/loss of one whole chromosome), or triploid 5 (additional pair of chromosomes). The distributions of euploid and aneuploid labels in the training, validation, and test sets are provided in Table 5.

#### 2.B.4. Sequence-level pregnancy outcomes

In the final dataset, 1,680 embryos were transferred to patients for pregnancy. Pregnancy outcome labels are subdivided by each survival stage: implantation, gestational sac, heartbeat and live birth. An unknown outcome refers to cases where a double embryo transfer is performed but only a single embryo progresses with the pregnancy, so that it is impossible to identify which of the two embryos survived. In total, 903 embryos have a positive implantation label, 557 negative, 220 unknown; 842 embryos had a gestational sac detected, 686 negative, 152 unknown; 757 had fetal heartbeat detected, 795 negative, 128 unknown; and 673 ultimately resulted in live birth 896 negative, 111 unknown. The distribution of pregnancy outcome labels is illustrated in Table 6.

## 3. Data collection and processing

The dataset consists of image sequences exported from the EmbryoScope time-lapse system and medical records retrieved from the clinic proprietary record software. Image pre-processing includes reading raw data, detecting unusable images using a convolutional neural network classifier, joining images with developmental stage and hour past insemination metadata, and writing sequences to hdf5 (4) format file, as described in Section 3.A. EHR pre-processing includes exporting tabular data from the clinic’s proprietary record-keeping software, applying basic type conversion and cleaning for consistency in manually-entered columns, and saving a subset of columns, to produce clean medical record features and outcome labels. Further details are described in Section 3.B. In this section we use word dataset in context of hdf5 files, to refer to ‘column’ of data. This is deliberate choice, to use language consistent with hdf5 documentation. We use “split(s)” to indicate subset of particular file with examples dedicated for either: training, validation or test purposes.

### 3.A. EmbryoScope data

The EmbryoScope device contains a Hoffman Modulation Contrast (HMC) microscope to capture images of individual embryos. Images are acquired every 10-15 minutes. At each round of acquisition, also referred to as “run”, the embryo is imaged at 7 focal planes with an increment of 15 micrometers. The embryo is contained in a conical well. The average number of runs per file after filtering is 499.3 (min. 273, max. 835). Which after filtering (see Section 4) corresponds to 476.7 (min. 128, max. 824) frames per sequence. Raw EmbryoScope data can be exported as SQL database files, which is the format retrieved for our dataset. This includes one image database file for each slide of embryos, as well as two databases of metadata: (i) patient information (PatientDB.fdb), and (ii) user annotations and machine environment properties (MainDB.fdb).

Within the image database files, an individual embryo is identified by combination of slide identifier and well number within the slide, and each image database file includes up to 12 embryos from the same treatment. Each file has a GENERAL table, with metadata such as time of insertion and measurements of the well environment, and an IMAGES table for the image sequences, where each row corresponds to a single monochromatic image of 500*×* 500 pixels in the format of a byte string which decodes to an image file. The images are indexed by (well number, run number, focal plane) and are associated with a raw time stamp of when the image was captured.

The initial image extraction consists of reading the image byte strings from the database files. In particular, the extraction procedure queries each database file for the IMAGES table, excluding calibration runs identified by run number *<* 1. We extract and save only the center focal plane of each imaging run, and we resize images to 250 *×* 250 pixels to save the disc space and reduce data loading and computational costs. Eventually, during training images are usually resized or cropped to 224 *×* 224, so we leave extra pixels for augmentation purposes. We store sequences in combined hdf5 chunked dataset, with chunk size equal to single image dimensions. To address individual images or sequences, we additionally store cumulative sums of sequence length in separate hdf5 dataset in the same file. We opt for single tensor solution for ease of management implied by uniform access pattern between different types of time series columns and little overhead, but may consider splitting sequences into separate datasets once read latency becomes a limiting factor. For all sequences, the time stamp of the first run corresponds to the approximate time of insemination, so we convert the time stamp to hours post insemination (“hpi”) and save it as dataset in the same hdf5 as the rest of data, with single vector format corresponding to images. For each well, we also retrieve development stage annotations from the MainDB.fdb file, join them with images using the raw time stamp, and save the annotations as corresponding vector of stage ids in the hdf5 dataset. It’s important to note that, at times, multiple annotations (usually two) may be assigned to a single image. This typically occurs when a stage transition happens between acquisition intervals. In cases of such annotation overlap, we resolve the conflict by favoring the more advanced stage. This approach may lead to inaccurate single-frame annotations for stages captured in just one frame, or errors where the more advanced stage is assigned to the frame preceding an unseen stage transition, rather than the one following it. However, we opt for simpler conflict resolution scheme considering total number of collisions is small: 47 individual collisions in final dataset. We then filter the images for valid quality as detailed in Section 4. In addition, we augment the hdf5 dataset with features and labels from the EHR. Each feature is individually stored as separate dataset with the sequence order. The final output is an hdf5 file containing image data, image-level metadata, and sequence-level features and labels for each embryo. Layouts and descriptions of derived and untypical columns can be found in Table 7. This overall image extraction is performed on the complete set of embryo ids which are valid according to the overall EHR filtering described in Section 4.A. We subsequently save a three different hdf5 files for each prediction task dataset. We split each dataset into train, validation and test set, with prior modification, by removing embryos which are filtered by task-specific criteria and augmented by data available only after certain point, like pre-transfer stimulation is only added to pregnancy outcomes prediction dataset (Section 2.B.4). We derive ploidy splits by individually filtering blastocyst formation splits, to ensure same patients are present in both datasets and models can be trained in multi-task fashion, or transferred from one task to another without data leak. Similarly pregnancy outcome splits are derived by filtering ploidy splits. **Note:** image-level development stage prediction dataset (Section 2.B.2) is split independently.

### 3.B. Clinical data

The clinical EHR data is accessed through a proprietary software API and can be exported as comma-separated values (CSV) file. It includes properties related to patient etiology, oocytes, ovarian stimulation, sperm measurements, endometrial preparation, and transfer, as well as the outcome labels described in Section 2.B. Table 8 provides a full list of the EHR features available, their definitions, and their possible values. For our prediction tasks, we focus on subsets of the full EHR features:

- **Patient etiology:** age, age of partner, demographics, hormonal measurements, antral follicle count (AFC), BMI, lifestyle factors (like smoking), medical, obstetric, and surgical histories.
- **Ovarian stimulation data:** ovarian stimulation protocol, stimulation priming, hormonal measurements, outcome measures (total oocytes and mature oocytes collected).
- **Oocyte data:** includes early or late oocyte maturation, whether the oocyte was cryopreserved, and fertilization method.
- **Genetic properties:** biopsy conclusiveness and subcategories of aneuploidy and monogenic disorder.
- **Sperm quality** sperm motility and concentration measurements before and after treatment
- **Endometrial preparation data:** endometrial preparation protocol, patient characteristics at transfer.
- **Transfer data:** number of embryos transferred, whether the embryo was transferred fresh or after cryopreservation, embryo age at time of transfer, in number of days, and transfer difficulty.

**Table 1.** Definitions of outcome labels assigned to each embryo.

| Name | Definition |
| --- | --- |
| Blastocyst formation | Positive if embryo reached any stage of blastulation (tSB, tB, tEB, tHB). |
| Ploidy | Positive (euploid) if biopsied embryo tissue shows 23 chromosome pairs. |
| Gestation | Positive if a gestational sac is detected. |
| Heartbeat | Positive if a fetal heartbeat is detected. |
| Live birth | Positive if the pregnancy successfully develops to live birth. |

**Table 2.** Blastocyst label distributions for each dataset split.

| Label | Train | Validation | Test |
| --- | --- | --- | --- |
| Positive | 5,860 | 2,082 | 2,021 |
| Negative | 3,059 | 885 | 869 |

**Table 3.** Definitions of development stage labels assigned to individual images and the stage label distributions for each dataset split.

| Stage | Definition | Train | Validation | Test |
| --- | --- | --- | --- | --- |
| tPB2 | second polar body appearance | 7,202 | 2,402 | 2,320 |
| tPNa | pronuclei appearance | 7,270 | 2,445 | 2,388 |
| tPNf | pronuclei fading | 7,778 | 2,540 | 2,564 |
| t2 | second cell appears | 7,787 | 2,562 | 2,563 |
| t3 | third cell appears | 5,720 | 1,920 | 1,914 |
| t4 | fourth cell appears | 7,356 | 2,425 | 2,417 |
| t5 | fifth cell appears | 6,471 | 2,179 | 2,173 |
| t6 | sixth cell appears | 6,599 | 2,177 | 2,169 |
| t7 | seventh cell appears | 6,330 | 2,099 | 2,016 |
| t8 | eight cell appears | 6,608 | 2,170 | 2,142 |
| t9 | ninth cell appears | 6,161 | 2,033 | 2,010 |
| tSC | start of compaction | 5,846 | 1,955 | 1,883 |
| tM | morula formation | 6,062 | 1,982 | 1,982 |
| tSB | blastulation start | 5,788 | 1,894 | 1,905 |
| tB | blastocyst formation | 4,670 | 1,500 | 1,498 |
| tEB | blastocyst expansion | 3,391 | 1,019 | 1,092 |
| tHB | blastocyst hatching | 1,357 | 425 | 405 |

**Table 4.** Definitions of development stage labels assigned to individual images and the stage label distributions for each dataset split after label filling.

| Stage | Train | Validation | Test |
| --- | --- | --- | --- |
| tPB2 | 143,672 | 49,606 | 43,588 |
| tPNa | 442,490 | 152,167 | 145,529 |
| tPNf | 106,368 | 34,275 | 34,170 |
| t2 | 310,770 | 109,461 | 102,951 |
| t3 | 124,918 | 41,900 | 48,357 |
| t4 | 339,530 | 116,512 | 116,989 |
| t5 | 178,569 | 56,188 | 65,932 |
| t6 | 190,754 | 60,561 | 67,462 |
| t7 | 198,372 | 69,732 | 65,237 |
| t8 | 325,070 | 106,228 | 103,975 |
| t9 | 384,439 | 126,038 | 131,670 |
| tSC | 256,064 | 87,799 | 77,832 |
| tM | 292,678 | 94,784 | 92,020 |
| tSB | 351,187 | 119,055 | 122,939 |
| tB | 152,144 | 52,421 | 48,278 |
| tEB | 147,833 | 44,740 | 52,145 |
| tHB | 56,711 | 17,787 | 17,131 |

**Table 5.** Ploidy label distributions for each dataset split.

| Label | Train | Validation | Test |
| --- | --- | --- | --- |
| Euploidy | 2,091 | 800 | 762 |
| Aneuploidy | 2,521 | 862 | 836 |

**Table 6.** Pregnancy label distributions for each dataset split.

| Task | Label | Train | Validation | Test |
| --- | --- | --- | --- | --- |
| Implantation | Positive | 524 | 193 | 186 |
|  | Negative | 318 | 123 | 116 |
|  | Unknown | 128 | 51 | 41 |
| Gestational sac | Positive | 489 | 180 | 173 |
|  | Negative | 392 | 157 | 137 |
|  | Unknown | 89 | 30 | 33 |
| Fetal heartbeat | Positive | 441 | 165 | 151 |
|  | Negative | 454 | 180 | 161 |
|  | Unknown | 75 | 22 | 31 |
| Live birth | Positive | 390 | 147 | 136 |
|  | Negative | 516 | 198 | 182 |
|  | Unknown | 64 | 22 | 25 |

**Table 7.** Metadata that is extracted from EmbryoScope data and stored with the final hdf5 files.

| Metadata | Description |
| --- | --- |
| 'patient_id' | Sequence-level patient identifier |
| 'well_id' | Sequence-level well identifier |
| 'cum_seq_len' | Sequence-level cumulative sequence length |
| 'run' | Image-level frame number |
| 'hpi' | Image-level hours post insemination |
| 'stage' | Image-level embryo stage from annotations (with label filling) |
| 'stage_no_fill' | Image-level embryo stage from annotations (without label filling) |
| 'image_label' | Image-level blastocyst indicator (derived from 'stage') |
| 'images' | Image at respective run |
| 'is_well_full' | Image-level binary flag (used for filtering) |
| 'sequence_label' | Sequence-level label indicating if sequence reached blastulation (for convenience) |
| other EHR features | Sequence-level values individually described in Table 8 |

**Table 8.**
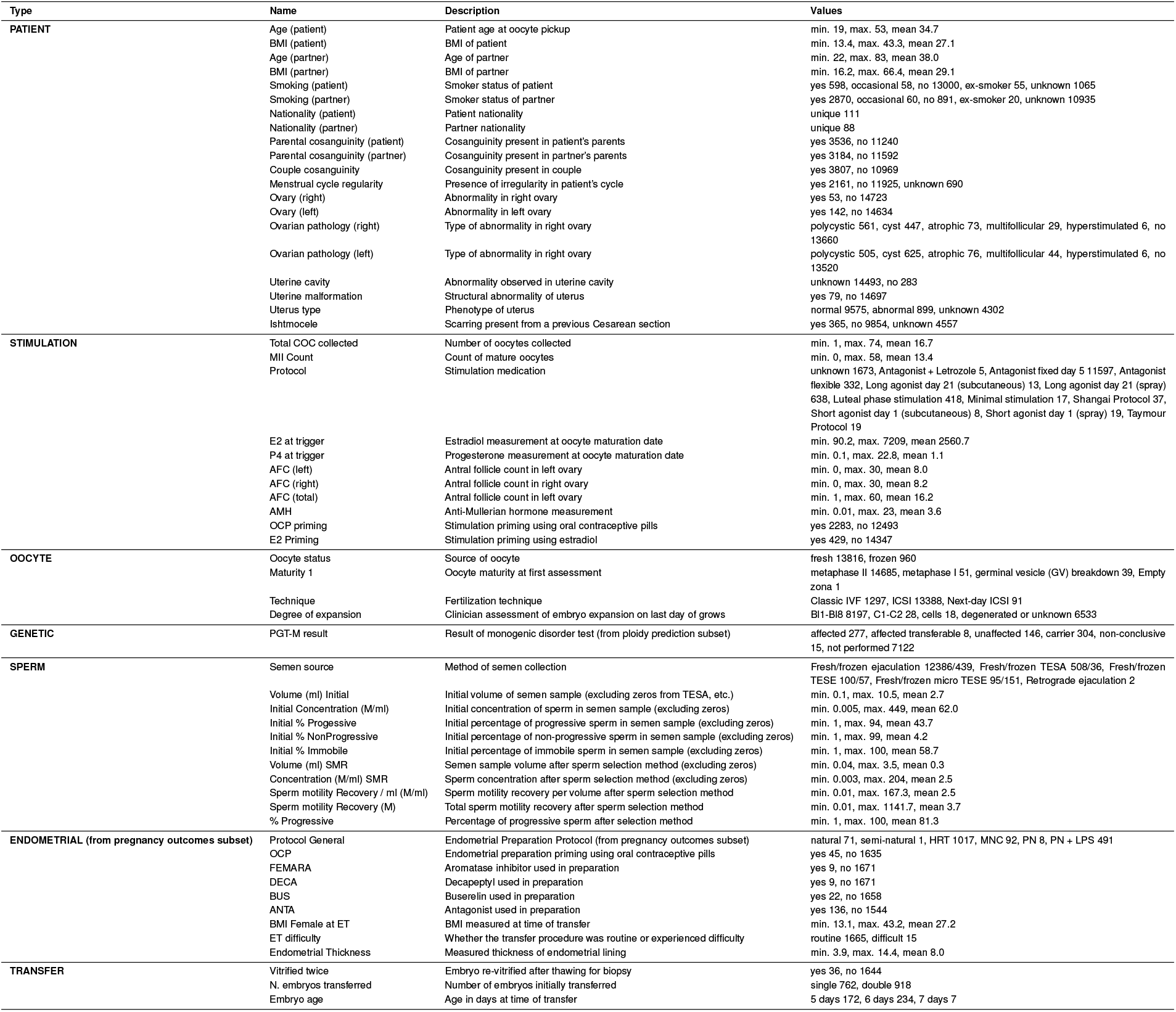
Clinical features extracted from the EHR data along with their definitions.

**Fig 1.**
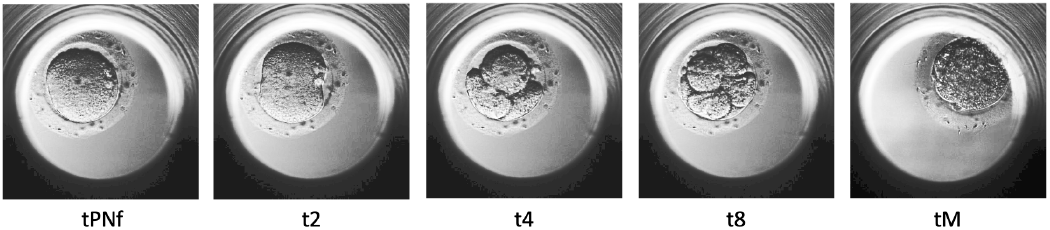
Microscopy images of embryos with corresponding developemnt stage labels.

**Fig 2.**
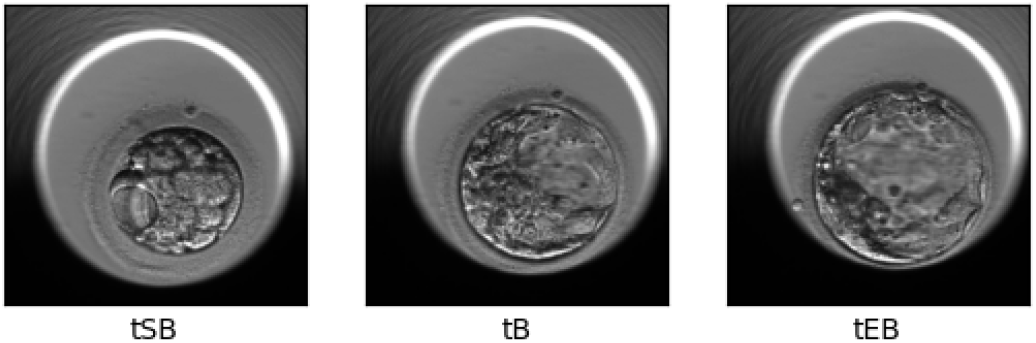
Examples of embryos in different stages of blastocyst formation.

## 4. Filtering

We filter samples based on both image sequences and EHR data. In Section 4.A, we describe the filtering procedure with the associated statistics at each step. In Section 4.B, we describe the process of training an image classifier to detect empty images and images with excessive artifacts.

### 4.A. Application of inclusion and exclusion criteria

The raw EHR record initially contains 40,392 (2,206 patients, 3,366 treatments) embryos with ART identifiers. We filtered this set based on both record integrity criteria and clinical exclusion criteria to obtain sequence-level blastocyst formation prediction dataset 2.B.1:

- First, we excluded 19,424 oocyte (43 patients, 97 treatments) records belonging to wells that had abnormal fertilization or didn’t fertilize at all.
- We then excluded 3,272 oocyte (205 patients, 505 treatments) records belonging to slides that were clinically not useful, such as those belonging to test slides, or slides
- which are duplicated because a new id was generated after re-inserting the slide after removing embryo for biopsy, or embryos were not incubated for long enough due to regulatory and changing clinical best practice reasons.
- Next, we linked the oocyte records with the extracted imaging sequences, and excluded 366 oocyte (38 patients, 59 treatments) records that did not have a corresponding .pdb file.
- We also excluded 233 oocytes (21 patient, 39 treatments) with corrupted (for unknown reason) image records.
- After aligning images with corresponding EHR records we employ our image classifier, to detect invalid images, which includes: lack of embryo in frame, excessive artifacts, like air bubble that obstructs visibility and other (refer to Section 4.B). We show samples of invalid images in Figure 3. We subsequently exclude continuous sequences of leading and trailing frames, marked as “invalid”.

**Fig 3.**
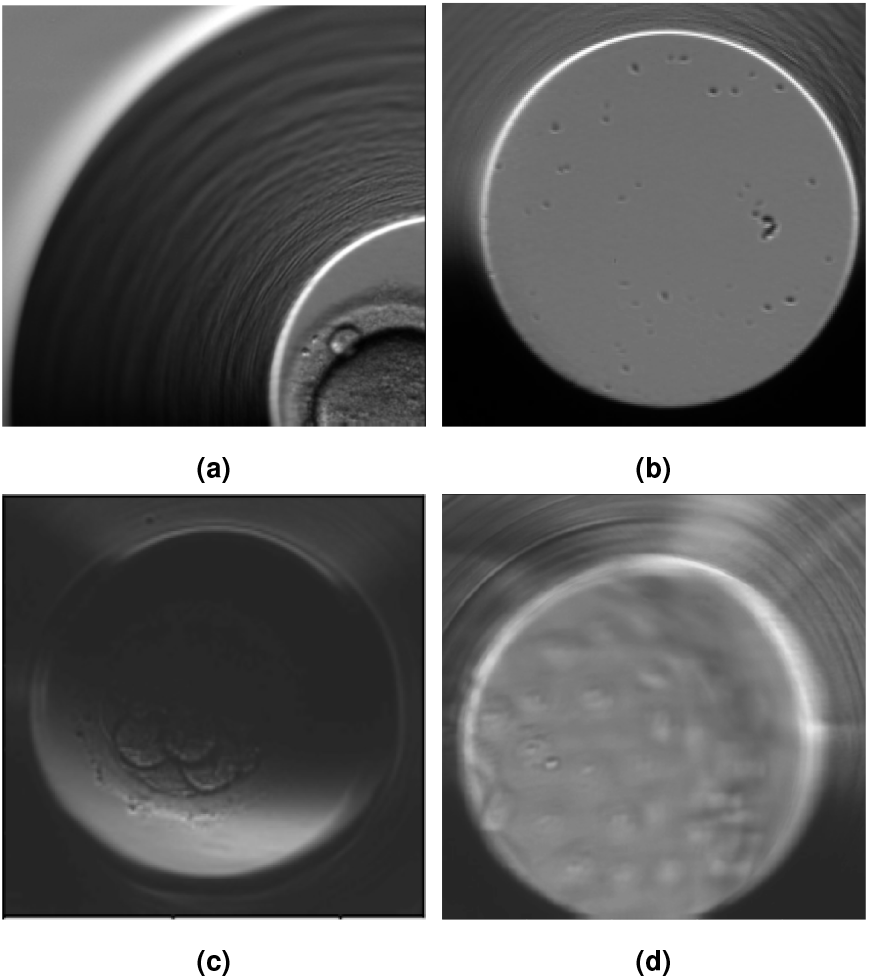
Examples of artifacts detected in the raw dataset: (a) a well which was misaligned with the camera, (b) an empty well, (c) An image which is excessively dark, and (d) a hatched blastocyst.

- Sequences containing 128 or fewer images after filtering are considered unusually short. This threshold was chosen based on the length of the shortest sequence that was deemed valid after manual review. Sequences that depict less than 96 hours of embryo development, based on the “hpi” feature of the last included image, are excluded as easy negatives. Additionally, we discard sequences with more than 15 marked images, after removing the leading and trailing images. The threshold for marked images was set at the 95th percentile, as the 96th percentile represented an abrupt increase, consisting of 30 empty images, leading to a disproportionate number of invalid observations. In total, this procedure excludes 2,321 embryos (89 patients, 166 treatments), amounting to 1,250,102 individual images.
- Additionally this process removes total of 1,019,404 images from the rest of 14,776 embryos (1,810 patients, 2,500 treatments).

For the image-level development stage dataset (Section 2.B.2), we exclude sequences with potentially incomplete information about embryo development stages. This includes sequences where embryo annotations start later than the t2 stage, indicating a significant gap in the annotation of early development stages. We also remove sequences that lack development stage annotations or show a mismatch between stage annotations and EHR records, particularly where blastulation labels are missing despite indications of blastocyst formation in the EHR. Additionally, we exclude sequences labeled with the tDead stage, as these may lack complete stage labeling due to the embryo’s non-viability for further transplantation. In total, this process removes 799 embryos (12 patients, 16 treatments) and 366,533 images.
For sequence-level genetic outcome prediction dataset 2.B.3, we apply additional filtering step to sequence-level blastocyst formation dataset, since we don’t necessarily want to rely on stage annotations. Final dataset excludes 6,876 embryos (160 patients, 290 treatments) that did not undergo biopsy, and additional 28 embryos (1 patient, 4 treatments) for which biopsy was inconclusive. Which gives a total of 7,872 embryos (1,807 patients, 2,480 treatments).

For the sequence-level pregnancy outcome prediction dataset 2.B.4 we further filtered genetic outcome prediction dataset, by including only embryos which were selected for transfer, which excludes 6059 embryos (607 patients, 1020 treatments). Out of those selected for transfer 72 embryos (9 patients, 17 treatments), from those cryopreserved, did not survive warming. For outcome labels we exclude 61 (35 patients, 40 treatments) embryos without full set of results for all four labels, stemming from failure to follow up. Final dataset contains 1,680 embryos (998 patients, 1,129 treatments), which includes 71 embryos (46 patients, 47 treatments) transferred fresh and 1,609 embryos (966 patients, 1,086 treatments) transferred after cryopreservation, here we observe the overlap between patients and treatments in fresh and cryo transfers.
**Note:** In some cases, pregnancy outcome labels cannot be unambiguously determined when double embryo transfer is performed, but only one survives. In such situations, it is unclear which embryo should receive a positive or negative label. However, if neither embryo progresses to the next pregnancy stage, the samples can again be labeled without ambiguity. Rather than excluding embryos with uncertain outcomes, we leave the decision to the specific learning scheme used.

### 4.B. Invalid image classification model

To detect valid images, we trained a convolutional neural network to detect (i) wells with no embryo, (ii) images where the well is shifted substantially out of the image frame, (iii) images with excessive lighting artifacts such as low contrast or blurring, and (iv) images of embryos that drifted or grew out of the camera view field. Images of wells which do not contain an embryo occur when the camera for a given well is turned on before the embryo is inserted in the well or after the well is removed (for biopsy, cryopreservation or growth medium change). Due to clinic practices, the image sequences for all wells in each slide begin when at least one embryo is inserted, and end when the last embryo is removed. A continuous sequence of empty images in the end of frame sequence appears when one or more embryos are removed earlier than the final embryo in the slide. Embryos may also be inserted later when classic IVF or next-day ICSI technique is used, as fertilization occurs in a separate medium before the embryo is transferred to the EmbryoScope well, resulting in a continuous sequence of leading empty images. Fig. 3 illustrates examples of images which are classified as “invalid”.

To train the classification network, we first created a dataset of 2,054 invalid and 11,325 valid image examples, identified through manual inspection. This dataset was then divided into training, validation, and test sets in a 60/20/20 ratio, with label stratification to ensure that each split maintains the original proportion of invalid to valid images.

**Note:** To preserve data integrity, we explicitly excluded all sequences used in training and/or evaluation of this model from the validation and test sets of the target dataset described in this report. This ensures that the validation and test results of downstream models are not influenced by any additional data purity.

We used these samples to train a ResNet-18 model with ImageNet weight initialization. We train model for 10 epochs, using Adam optimizer with a learning rate of 0.001. We use linear learning rate schedule with 1000 steps of warm up. To improve performance, we apply data augmentation techniques including random horizontal and vertical flip, and random cropping from 250 *×* 250 to 224 *×* 224. We employ cropping to account for potential small shifts in embryo position, but not enough to introduce change of the label.

## 5. Conclusion

The ART Dataset is a real-world multimodal clinical dataset which combines detailed clinical features with high-resolution image time-series, to provide a rich source of information for analyzing fertility treatment outcomes and training machine learning models to automate labeling and clinical prediction tasks.

## Data Availability

Data presented in the study is private and can only be made available to collaborators in the UAE, due to local regulation, upon reasonable request and ethical approval

## Footnotes

* https://www.vitrolife.com/products/time-lapse-systems/embryoscope-8-time-lapse-system/

